# Cell-based culture of SARS-CoV-2 informs infectivity and safe de-isolation assessments during COVID-19

**DOI:** 10.1101/2020.07.14.20153981

**Authors:** K Basile, K McPhie, I Carter, S Alderson, H Rahman, L Donovan, S Kumar, T Tran, D Ko, T Sivaruban, C Ngo, C Toi, MV O’Sullivan, V Sintchenko, S C-A Chen, S Maddocks, DE Dwyer, J Kok

## Abstract

**Background:** The detection of SARS-CoV-2 by real-time polymerase chain reaction (PCR) in respiratory samples collected from persons recovered from COVID-19 does not necessarily indicate shedding of infective virions. By contrast, the isolation of SARS-CoV-2 using cell-based culture likely indicates infectivity, but there are limited data on the correlation between SARS-CoV-2 culture and PCR. Here we review our experience using SARS-CoV-2 culture to determine infectivity and safe de-isolation of COVID-19 patients.

**Methods:** 195 patients with diverse severity of COVID-19 were tested (outpatients [n=178]), inpatients [n=12] and ICU [n=5]). SARS-CoV-2 PCR positive samples were cultured in Vero C1008 cells and inspected daily for cytopathic effect (CPE). SARS-CoV-2-induced CPE was confirmed by PCR of culture supernatant. Where no CPE was documented, PCR was performed on day four to confirm absence of virus replication. Cycle threshold (Ct) values of the day four PCR (Ct_culture_) and the PCR of the original clinical sample (Ct_sample_) were compared, and positive cultures were defined as a Ct_sample_ - Ct_culture_ value of ≥3.

**Findings:** Of 234 samples collected, 228 (97%) were from the upper respiratory tract. SARS-CoV-2 was only successfully isolated from samples with Ct_sample_ values <32, including in 28/181 (15%), 19/42 (45%) and 9/11 samples (82%) collected from outpatients, inpatients and ICU patients, respectively. The mean duration from symptom onset to culture positivity was 4.5 days (range 0-18 days). SARS-CoV-2 was significantly more likely to be isolated from samples collected from inpatients (p<0.001) and ICU patients (p<0.0001) compared with outpatients, and in samples with lower Ct_sample_ values.

**Conclusion:** SARS-CoV-2 culture may be used as a surrogate marker for infectivity and inform de-isolation protocols.

## Introduction

In late December 2019, a cluster of pneumonia cases of unknown aetiology were reported in Hubei Province, China^1^. The causative agent, a novel beta-coronavirus, now known as SARS-CoV-2, was isolated on January 7, 2020.^2^ The genetic sequence of SARS-CoV-2 was shared on January 10, 2020, enabling the rapid development of diagnostic assays to test persons with suspected COVID-19 (the disease state caused by SARS-CoV-2) to be available within a month of the outbreaks’ notification to the World Health Organization (WHO).^3^

Effective control of SARS-CoV-2 transmission requires widespread testing, isolation of infected persons and contact tracing, particularly in the absence of effective SARS-CoV-2-specific antivirals and vaccines. Nucleic acid testing such as real-time reverse transcriptase polymerase chain reaction (PCR) is the predominant laboratory diagnostic method to confirm infection, however, detection of SARS-CoV-2 RNA does not necessarily indicate viable and transmissible virus. Apart from its role in the initial identification of SARS-CoV-2, the diagnostic value of culture in the routine management of COVID-19 is limited by requirements of virus isolation to be performed under physical containment laboratory level 3 (PC3) biosafety conditions,^4^ technical expertise, turnaround times and costs. However, the ability to culture SARS-CoV-2 has important roles in providing adequate positive control material for the development, validation, evaluation and quality assurance of SARS-CoV-2 diagnostic assays, supporting the development of vaccines and therapeutic agents, and enabling research into viral virulence and transmission.

As the pandemic has evolved it has become clear that there are a proportion of patients where SARS-CoV-2 RNA remains detectable for many days to weeks.^5^ This has significant implications for the timing of release from quarantine and is particularly important for those individuals who reside in, or work with, high-risk population groups (such as healthcare workers (HCWs), aged care facilities, correctional centres, Indigenous communities, immunosuppressed). Persistent SARS-CoV-2 RNA detection also has implications for patient isolation and bed management, the safe use of non-invasive ventilation and ongoing requirements for HCWs to wear appropriate personal protective equipment (PPE) when caring for such patients. However, the detection of SARS-CoV-2 RNA may not always indicate the presence or shedding of viable virus with replicative capacity and, by implication, transmissibility.^6^

Herein, we investigate the correlation between SARS-CoV-2 culture and PCR results from respiratory tract samples collected from persons with COVID-19 to guide the safe de-isolation of those with persistently positive PCR.

## Methods

Results of respiratory tract samples received between March 1 and April 22, 2020 for SARS-CoV-2 PCR that were also inoculated into continuous cell culture were analysed. Samples were inoculated into cell culture as part of (i) routine laboratory protocol at the start of the outbreak; (ii) if collected from patients admitted to the intensive care unit (ICU), and (iii) if requested by attending physicians (usually due to persistent PCR positivity in an individual awaiting release from quarantine) or public health units as part of outbreak investigations.

### Sample types

Samples received for SARS-CoV-2 PCR included upper respiratory tract (URT; nasopharyngeal swabs [NPS] and combined nose and throat swabs [NTS]) and lower respiratory tract (LRT; sputum, endotracheal aspirate [ETA], non-bronchoscopic bronchoalveolar lavage [nbBAL]) samples. URT swabs were received in 1-3 mL of viral or universal transport media (UTM), Amies transport media, or dry; transported to the laboratory, stored at +4°C and tested within 12 hours of specimen receipt.

### SARS-CoV-2 nucleic acid detection in respiratory tract samples

RNA was extracted from 200μL of respiratory tract sample using the MagNA Pure 96 instrument’s Viral NA Small Volume Kit (Roche Diagnostics GmbH, Mannheim, Germany) following the “Pathogen Universal Protocol” (elution volume, 100μL) as per the manufacturers’ instructions. SARS-CoV-2 nucleic acid amplification and detection by RT-PCR were performed using the LightCycler® 480 II System (Roche Diagnostics, Rotkreuz, Switzerland). The primers and probes, mastermix composition and cycling conditions used for the in-house developed assays are outlined in Table 2 (Supplementary). Each PCR assay included a RNAse-free water negative control, positive control of a 10^−3^ dilution of a SARS-CoV-2-positive cell culture supernatant and a human beta-globin gene (hBGL) internal control. Cycle threshold (Ct) values of positive samples were recorded.

SARS-CoV-2 gene targets for PCR included E, RdRp, N, M, and ORF1ab for samples from ICU patients; and between one and four of E, RdRp, N and Orf1ab for all other samples. Shortages of PCR reagents and consumables, both globally and in Australia, influenced the targets tested. The total number of samples tested per target were: one (n=24), two (n=115), three (n=22), four (n=62) and five targets (n=11).

### SARS-CoV-2 culture of respiratory tract samples

Samples where SARS-CoV-2 RNA was detected were inoculated into cell culture the same day or were stored at +4°C for inoculation the following day. Vero C1008 (Vero 76, clone E6, Vero E6 [ECACC 85020206]) cells were seeded at 1-3 × 10^4^ cells/cm^2^ with Dulbecco’s minimal essential medium (DMEM, LONZA, Alpharetta, GA, USA) supplemented with 9% foetal bovine serum (FBS, HyClone, Cytiva, Sydney, Australia) in 25cm^2^ cell culture flasks or Costar® 24-well clear TC-treated multiple well plates (Corning®, Corning, NY, USA). The media was changed within 12 hours for inoculation media containing 1% FBS with the addition of penicillin, streptomycin and amphotericin B deoxycholate to prevent microbial overgrowth and then inoculated with 500μL (cell culture flasks) or 200μL (24-well plates) of clinical sample. Plates were sealed with AeraSeal™ Film (Excel Scientific, Inc., Victorville, CA, USA) to minimise evaporation, spillage, and well-to-well cross-contamination. The inoculated cultures were incubated at 37° C in 5% CO_2_ for 5 days (day 0 to 4). Cell cultures were observed daily for cytopathic effect (CPE) and 200μL of viral culture supernatant was collected on days 1, 2, 3 and 4 for the ICU cohort and on day 4 for all other samples. Day four was chosen for terminal sampling as this was the optimal time for reading the endpoint as determined by the TCID_50_ assay (see below). Additionally, with no media change and using the upper range concentration of amphotericin B (2.5μg/mL), cell monolayer breakdown was observed from day five onwards in some cultures. Routine mycoplasma testing was performed to exclude mycoplasma contamination of the cell line and all culture work was undertaken in a physical containment laboratory level 3 (PC3) biosafety conditions.^4^

Where CPE was observed, SARS-CoV-2 replication was confirmed by PCR. CPE, when observed, generally occurred within 2-4 days post-inoculation. Terminal sampling of culture supernatant was performed on day four using PCR as CPE may not be observed despite active viral replication, as seen with SARS-CoV-1.^7^

### SARS-CoV-2 nucleic acid detection in viral culture supernatants

Viral RNA was extracted from 200μL of viral culture supernatant using the EZ1 Advanced XL platform and EZ1 Virus Mini Kit v2.0 (Qiagen, Valencia, CA, USA) according to the manufacturers’ instructions. SARS-CoV-2-specific PCR was then performed as indicated above, and Ct values recorded.

### Determination of TCID_50_

A primary isolate, SARS-CoV-2 /01/Human/NSW Australia|EPI_ISL_407893|2020-01-24 (GISAID), was titrated in serial one log dilutions (from 10^−1^ to 10^−8^) to obtain a 50% tissue culture infective dose (TCID_50_) using 24-well culture plates of Vero E6 cells. The plates were observed daily for CPE using an inverted optical microscope for five days, and wells were considered positive if any characteristic altered cell foci were seen (Figure 4, Supplementary). The end-point titres were calculated according to the Reed & Muench method^8^ based on four replicates for titration. Using this method, the infectious virus titre of our SARS-CoV-2 stock was 1 × 10^6.5^ infectious particles per 200μL.

### Determination of virus replication and Ct value cut-off for virus viability in cell culture supernatant

To clarify the significance of the PCR Ct value of cell culture supernatant in the absence of CPE, a dilutional series of virus stock was run (as per TCID_50_) and SARS-CoV-2 RNA detection by PCR targeting six genes (E, RdRp, M, N, ORF1ab and ORF1b) at each dilution was performed (Figure 1). This allowed the limit of detection of viral culture (TCID_50_) to be compared to the limit of detection of the various PCR gene targets and demonstrated that the highest Ct value reflective of non-viable virus was 37.29 with the ORF1b gene target. Using the same targets, the dilutional series also indicated a change in 1 log dilution of virus concentration correlated with a change in Ct value of approximately ±3. This was confirmed by a real time quantitative PCR assay demonstrating that a 1 log difference in viral load corresponded to a change in Ct value of 3.44 ± 0.16. This assay also demonstrated that a Ct value of 34 correlated with a viral load of approximately 100 copies/μL, and a Ct value of 37 with 10 copies/μL, a difference of 1 log (data not shown).

**Figure 1:**
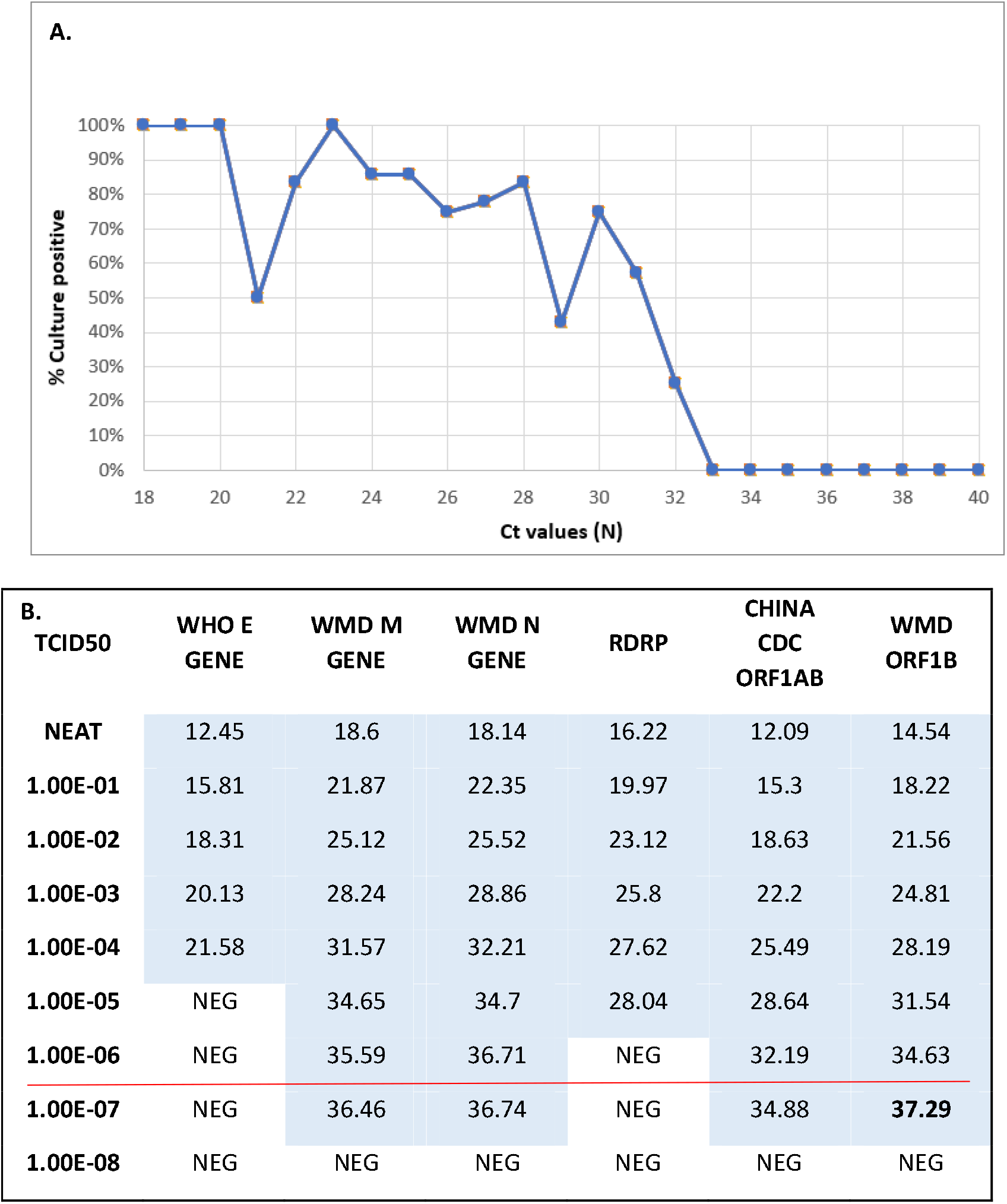
PCR Ct cut-off value for non-viability of clinical samples and for determination of culture positive versus negative in the absence of CPE. **A. Culture positivity rate by clinical sample PCR Ct value:** No clinical samples with PCR Ct values >32 resulted in positive viral cultures. Safety margin of 1log viral load less than this and concordant with B. was 37. **B: Culture supernatant PCR Ct values by gene target for TCID**_50_ **dilutions:** indicates differential sensitivity of PCR depending on target gene. Red line at 10^−6.5^ indicates limit of detection by TCID_50_. Ct value for ORF1b target of 37.29 (bold) chosen as highest Ct value representative of non-viable virus.

A positive viral culture was thereafter defined as (i) CPE visualised and SARS-CoV-2 RNA detected from cell culture supernatant, or (ii) no CPE visualised but a decrease in Ct values between the Ct of the original clinical sample PCR (Ct_sample_) and the terminal culture (day four) supernatant PCR (Ct_culture_) of ≥3 (equivalent to a 1 log increase in virus quantity) i.e. Ct_sample_ - Ct_culture_ ≥3 = culture positive. Conversely, a negative viral culture was defined as (i) CPE not visualised, and no SARS-CoV-2 RNA detected from the cell culture supernatant, or (ii) no CPE visualised in culture and Ct_sample_ - Ct_culture_ <3. Where SARS-CoV-2 RNA was detected, we surmise that Ct_sample_ minus Ct_culture_ <3 was due to residual inoculated clinical sample and not replicating virus. This is supported by the initial rise in Ct_culture_ on day 1 compared to Ct_sample_ on day 0 (Figure 2). In addition, no viral replication was identified with blind passaging of the first 20 cultures without CPE and Ct_sample_ - Ct_culture_ <3, indicating the absence of viable virus. By contrast, SARS-CoV-2 replication was demonstrated by CPE and PCR in subsequent passaging of samples without initial CPE but where Ct_sample_ minus Ct_culture_ was ≥3.

**Figure 2:**
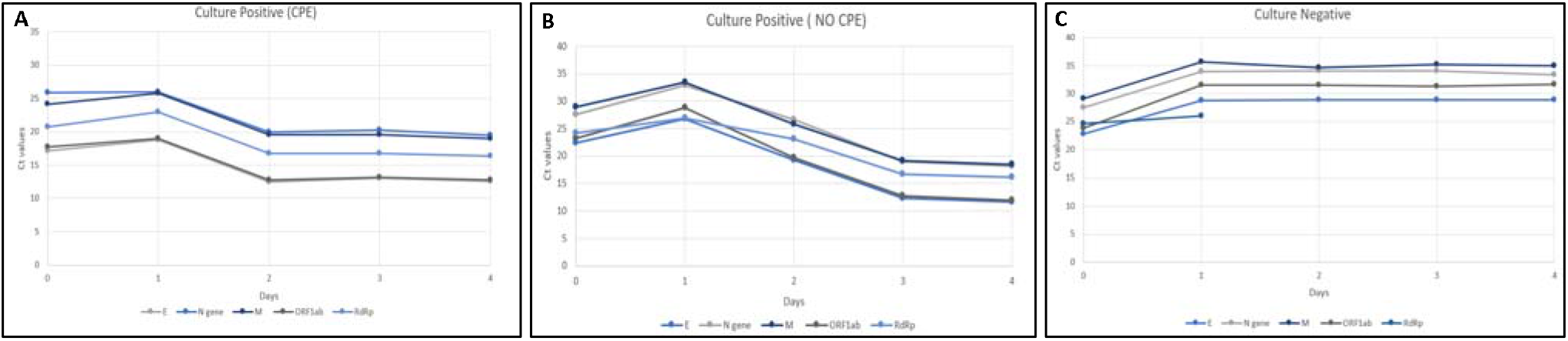
Representative samples from ICU cohort demonstrating target-dependent change in Ct values over four days of culture in **A**. a positive culture *with* CPE, **B**. a positive culture *wthout* CPE but Ct_sample_ - Ct_culture_ ≥3, and C. a negative culture (no CPE, PCR positive but Ct_sample_ - Ct_culture_ <3). Target genes: E, RdRp, M, N and ORF1ab.

### Statistical analyses

The Welch’s t-test using a two-tailed hypothesis with 95% confidence intervals (95% CI) was used to compare the mean Ct values of URT and LRT samples (Figure 6, Supplementary); of culture positive (with CPE), culture positive (without CPE) and culture negative samples (Figure 5, Supplementary), and to determine the mean Ct value of the N gene in each week of culture (Figure 3). Fisher’s exact test was used to determine significance in culture positivity between different clinical cohorts with 95% CI, (Table 1) and the difference between the culture positive samples per week (Figure 3).

**Table 1:** Patient demographics and sample characteristics.

|  | ICU | Inpatients<br>(non-ICU) | Outpatients<br>(Non-admitted) | Total |
| --- | --- | --- | --- | --- |
| **Number of patients** | 5 | 12 | 178 | 195 |
| **Average age (Range) yrs** | 58 (36-76) | 43 (22-78) | 40 (18-71) | 40 (8-78) |
| **Male gender** | 3 (60%) | 8 (70%) | 136 (77%) | 147 (75.4%) |
| **Number of samples** | 11 | 42 | 181 | 234 |
| **URT** | 6 | 40 | 181 | 227 (97%) |
| **LRT** | 5 | 2 | 0 | 7 (3%) |
| **Average days from symptom onset to sample collection (range)** | 7 (0-20) | 7 (1-16) | 12 (0-29) ** | 11 (0-29) |
| **Culture Positive & CPE** | 6 (55%) | 17 (40%) | 23 (12%) | 46 (20%) |
| **Culture Positive & no CPE** | 3 (27%) | 2 (5%) | 5 (3%) | 10 (4%) |
| **TOTAL Culture Positive** | 9 (82%) | 19 (45%) | 28 (15%) | 56 (24%) |
| **TOTAL Culture Negative** | 2 (18%) | 23 (55%) | 153 (85%) | 178 (76%) |

\*\* Time of symptom onset was unavailable for 56 samples collected from outpatients as part of contact investigations who were either asymptomatic or had no symptom onset date recorded.

**Figure 3:**
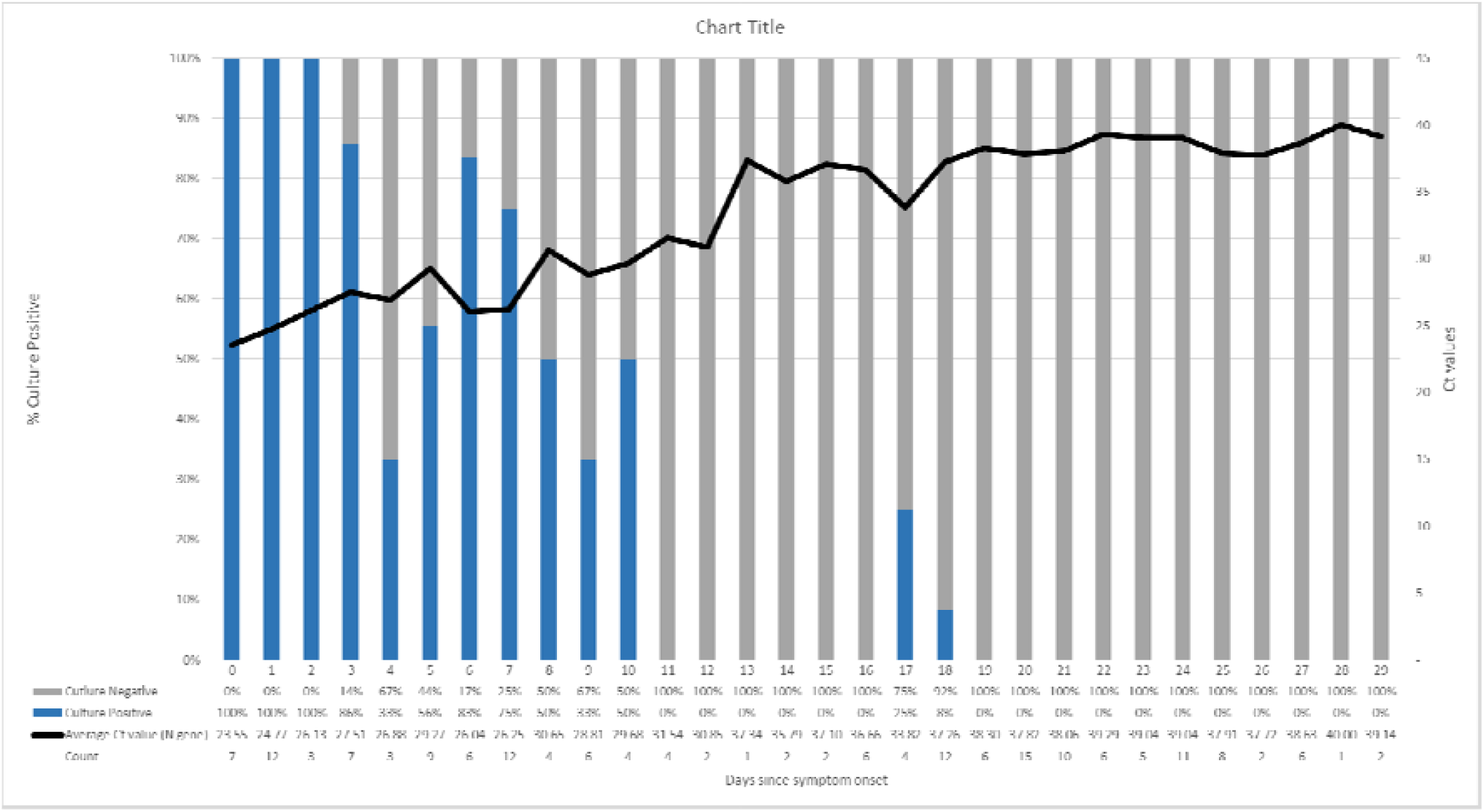
Comparison of Culture Positive and Negative samples and Days since symptom onset with the clinical sample N gene Ct value. ** Note: This figure excludes 56 samples where date of symptom onset was unknown (patient asymptomatic, or the date of symptom onset not recorded).

The Mann-Whitney U test using two-tailed hypothesis (95% CI) was used to compare the date of symptom onset in culture positive and negative patients (Figure 7, Supplementary) and the comparison of the median Ct value of the N gene in each week (Figure 3). *p*-values <0.05 were considered statistically significant.

## Results

A total of 234 samples from 195 persons with SARS-CoV-2 infection were included: 11 samples (5%) from five patients admitted to ICU, 42 (18%) from 12 inpatients that did not require ICU admission, and 181 (77%) from 178 outpatients (Table 1). Of the clinical samples tested, 228 (97%) were collected from the URT. The overall culture positivity rate was 56 (24%) and culture was significantly more likely to be positive in ICU patients compared with other inpatients (82% vs 45%, *p*=0.04) or outpatients (82% vs 15%, *p*<0.001); and also significantly more likely to be positive in samples from inpatients compared to outpatients (45% vs 15%, *p*<0.001) (Table 1).

CPE was observed and SARS-CoV-2 RNA was detected in cell culture supernatant in 46 (20%) samples. In 10 (4%) samples, no CPE was observed, but fulfilled the Ct_sample_ - Ct_culture_ ≥3 criteria to define positivity, and the remaining 178 (76%) were culture negative. Whole genome sequencing was performed on the ten samples deemed culture positive with no CPE and the lack of CPE was not associated with any SARS-CoV-2 lineage or isolated from any epidemiologically linked cluster (data not shown).^9^ Mean Ct values of the N gene were significantly lower in CPE-positive cultures when compared to both CPE-negative but PCR-positive cultures (25.01 vs 27.75, *p* <0.001) and to negative cultures (25.01 vs 36.87, *p* <0.001) (Figure 5, supplementary).

The highest Ct value in a clinical sample that was successfully cultured was 32 with the N gene target. Based on this result and the Ct cut-off value of 37 determined by PCR of TCID_50_ dilutions, and incorporating a 1 log margin of error, we were confident that any clinical sample with a Ct value of ≥37 was not indicative of viable virus (Figure 1).

Serial sampling of ICU patients and testing using multiple gene targets demonstrated the variability in Ct values when SARS-CoV-2 RNA was detected, with the E gene generally giving the lowest Ct values and N gene the highest (Figure 2) and (Figure 6, supplementary) in this cohort. There was consistent correlation over time between targets and concordance with the selected definition for culture positivity (Figure 2).

Whilst one patient continued to be culture positive to day 18 post-symptom onset, no others were positive beyond 10 days after symptom onset (Figure 3). The mean Ct value across all cultures increased as time from symptom onset increased (Figure 3). Using the N-gene as a representative target, cultures were significantly more likely to be positive from samples collected within the first week after symptom onset when compared to the second week (80% vs 45%, *p*=0.002), and from samples collected in the second week compared to the third week (45% vs 4%, *p*<0.001) (Figure 3).

Respiratory tract sample PCR Ct values of the most frequently used target (the N gene) ranged from 17.56 to 40, mean=34.16, IQR (29.21 –39.37). The mean Ct value of the N gene PCR was significantly lower in LRT compared with URT samples (26.76 vs 34.41, *p*<0.001) (Figure 6, supplementary). No significant difference was observed for the other targets.

In clinical samples where SARS-CoV-2 was isolated, the mean duration between symptom onset and sample collection was 4.5 days (median 3.5, range 0-18 days). By contrast, the mean duration between symptom onset and sample collection in culture negative samples was 20 days (median 18.4, range 3-29 days; *p*<0.001) (Figure 7, Supplementary).

## Discussion

Detection of SARS-CoV-2 RNA has been the primary diagnostic method to confirm COVID-19 disease. As an indicator of infectivity, the absence of detectable SARS-CoV-2 RNA in respiratory tract samples is utilised in many jurisdictions as a requirement for release of persons with COVID-19 from quarantine.^10^ In Australia, there has been a requirement that infected persons from high-risk settings (including but not limited to HCWs) have two consecutive negative respiratory tract SARS-CoV-2 PCRs taken at least 24 hours apart before being allowed to return to such settings.^11^ Whilst there are publications that have reported persistent or prolonged PCR positive results in recovered persons,^12,13^ few have provided robust data to support that they are no longer infectious. Furthermore, there is substantial variability in the duration of PCR positivity (up to 28 days after symptom onset),^12^ and this has resulted in individuals (including HCWs) being isolated for prolonged periods given the potential risks of virus transmission. As the detection of SARS-CoV-2 RNA in respiratory samples does not necessarily indicate viable infectious virions, we examined the correlation between the Ct values of PCR-positive samples and viral culture, a surrogate marker for infectivity. The Australian National Guidelines for Public Health Units also recommends that viral culture results be considered when deciding on the release of persons with COVID-19 from quarantine.^11^

We successfully isolated SARS-CoV-2 from upper and lower respiratory tract samples collected from persons with acute COVID-19 and correlated the yield of cell culture with PCR Ct values. Similar to other reports.^14,15^ We demonstrate that the threshold PCR Ct value where cell culture will be successful is less than 32 for the N gene target. However, this Ct value is specific for our laboratory PCR and we would recommend that diagnostic laboratories correlate their Ct values with culture results due to varying analytical performance of different SARS-CoV-2 PCR assays.^16^

The cell culture method described herein relies not only on detection of CPE but incorporates PCR testing. We demonstrate that virus with replicative capacity can be isolated and detected in the absence of CPE by correlating the Ct values of cell culture supernatant with the Ct values of the original clinical samples where SARS-CoV-2 was detected. This effect wasn’t due to a known SARS-CoV-2 genotype, however further genetic analysis may be warranted. This provides a more objective and sensitive measure of virus isolation as the detection of CPE can be subtle or absent due to low levels of virus that result in a delayed rise in virus load beyond day four in the cell lines used. This may assist in improving workflows in laboratories processing large numbers of cell cultures or those with reduced capacity to detect CPE. A culture supernatant Ct value cut-off >32 in our laboratory correlates with non-viable virus; and that in the absence of CPE, a Ct_sample_ - Ct_culture_ value <3 also indicates an absence of viral replication. This suggests that these individuals could be safely released from isolation.

We have demonstrated that the yield of culture is improved when samples are collected as close to the date of symptom onset as possible, and virus isolation is more likely in patients with more severe illness requiring admission to hospital or the ICU compared to outpatients. A previous study determined that virus may be isolated up to six days prior to symptom onset,^17^ but the samples collected in our study did not include such a cohort. Given the turnaround times, virus isolation of samples collected from persons in the pre-symptomatic stage would not be useful in a meaningful timeframe to guide clinical management, but would provide laboratory evidence of pre-symptomatic or asymptomatic SARS-CoV-2 transmission. We showed culture positivity out to 18 days post-symptom onset in one person, which is ten days longer than previous findings from Singapore.^15^

## Limitations

In the present study, we were not able to test all samples or cell culture supernatants with all five SARS-CoV-2 gene targets (E, RdRp, M, N and ORF1ab) due to reagent shortages. Nevertheless, we hypothesize that the findings using the N target would be similar for the other targets as a similar pattern was observed where testing of multiple targets were performed (Figure 2). Nucleic acid extraction from the primary sample and cell culture supernatant was performed on the MagNA Pure 96 and EZ1 Advanced XL instruments, respectively. Previous evaluations of automated nucleic acid extraction instruments for non-SARS-CoV-2 respiratory viruses showed superiority of the EZ1 Advanced XL compared to that of the MagNA Pure 96,^18^ but there is limited data on the performance of extraction instruments for SARS-CoV-2. We were unable to ascertain if different swab types or UTM affected either PCR or culture results. This study also did not investigate why certain individuals shed SARS-CoV-2 RNA for longer periods compared to others, nor were we able to determine whether there were any factors such as immunosuppression that may have explained prolonged viral shedding, and further research is recommended to address these questions.

## Conclusion

In summary, viral culture may assist clinicians to determine the safety of releasing from isolation individuals who have recovered clinically but remain persistently or intermittently positive SARS-CoV-2 by PCR. Whilst access to virus culture can differ in clinical diagnostic laboratories, this capacity adds important insights not provided by molecular methods and should be maintained in reference laboratories to guide clinical management and de-isolation.

## Data Availability

Data set not attached

## Author Contributions

**Literature search:** Basile K, Maddocks S and Kok J

**Study design:** Basile K, McPhie K, Carter I, Maddocks S and Kok J

**Figures:** Basile K and Maddocks S

**Data collection:** Basile K, McPhie K, Carter I, Alderson S, Rahman H, Donovan L, Kumar S, T Tran T, D Ko D, Sivaruban T, Ngo C and C Toi

**Data Analysis:** Basile K, Maddocks S and Kok J

**Data Interpretation/;** Basile K, Maddocks S and Kok J

**First Draft:** Basile K

**Editing and final draft:** Basile K, O’Sullivan MV, Sintchenko V, Chen SC-A, Maddocks S, Dwyer DE, and J Kok

## Ethics statement

Clinical specimens were processed at the NSW Health Pathology-Institute of Clinical Pathology and Medical Research (ICPMR) for standard of care diagnostic purposes and deemed not research. A non-research determination for this project was granted by the Health Protection NSW as it was a designated communicable disease control activity.

## Funding statement

This study was supported by the Prevention Research Support Program funded by the New South Wales Ministry of Health and the National Health and Medical Research Council (APPRISE 1116530). The funders had no role in the study design, data collection, data analysis and interpretation, or writing of the report. The corresponding author had full access to study data and final responsibility for the decision to submit for publication.

## Conflict of interest

None declared

## Acknowledgments

Jimmy Ng^1^ and Basel Suliman^1^

## Notes

### Competing Interest Statement

The authors have declared no competing interest.

## References

1. World Health Organization (WHO). Pneumonia of unknown cause – China. Geneva: WHO; 2020. Available from: https://www.who.int/csr/don/05-january-2020-pneumonia-of-unkown-cause-china/en/

2. The 2019-nCoV Outbreak Joint Field Epidemiology Investigation Team, Qun Li. An Outbreak of NCIP (2019-nCoV) Infection in China — Wuhan, Hubei Province, 2019-2020[J]. China CDC Weekly, 2020, 2(5): 79–80. Doi: 10.46234/ccdcw2020.022

3. https://www.who.int/docs/default-source/coronaviruse/situation-reports/20200121-sitrep-1-2019-ncov.pdf?sfvrsn=20a99c10_4

4. World Health Organization (WHO). Laboratory biosafety guidance related to coronavirus disease (COVID-19). WHO/WPE/GIH/2020.3. Available from: https://www.who.int/publications/i/item/laboratory-biosafety-guidance-related-to-coronavirus-disease-(covid-19)

5. Li N, Wang X, Lv T. Prolonged SARS-CoV-2 RNA shedding: Not a rare phenomenon [published online ahead of print, 2020 Apr 29]. J Med Virol. 2020;10.1002/jmv.25952. doi:10.1002/jmv.25952

6. Burkardt HJ. Standardization and quality control of PCR analyses. Clin Chem Lab Med. 2000;38(2):87–91. Doi:10.1515/CCLM.2000.014

7. Kaye M, Druce J, Tran T, et al. SARS-associated coronavirus replication in cell lines. Emerg Infect Dis 2006; 12:128–133.

8. Reed, L.J., Muench, H., 1938. A simple method of estimating fifty per cent endpoint. Am. J. Hyg. 27, 493–497.

9. Rockett, R.J., Arnott, A., Lam, C. et al. Revealing COVID-19 transmission in Australia by SARS-CoV-2 genome sequencing and agent-based modeling. Nat Med (2020). https://doi.org/10.1038/s41591-020-1000-7

10. Tay JY, Lim PL, Marimuthu K, et al. De-isolating Coronavirus Disease 2019 suspected cases: a continuing challenge. Clin Infect Dis 2020 Feb 26; ciaa179. doi: 10.1093/cid/ciaa179. Online ahead of print.

11. Communicable Diseases Network Australia (CDNA) and Pubic Health Laboratory Network (PHLN) joint statement. Revisions to the Australian criteria for release from isolation and return to high-risk settings for persons recovering from COVID-19. Available at: https://www1.health.gov.au/internet/main/publishing.nsf/Content/cdna-song-novel-coronavirus.htm

12. Wölfel R, Corman VM, Guggemos W, et al. Virological assessment of hospitalized patients with COVID-2019. Nature 2020; 581:465–469.

13. Gombar S, Chang M, Hogan CA, et al. Persistent detection of SARS-CoV-2 RNA in patients and healthcare workers with COVID-19. J Clin Virol 2020; 129: 104477.

14. La Scola B, Le Bideau M, Andreani J, et al. Eur J Clin Microbiol Infect Dis 2020; 39(6): 1059–61.

15. National Centre for Infectious Diseases and the Chapter of Infectious Diseases Physicians, Academy of Medicine, Singapore – 23 May 2020. Period of Infectivity to inform strategies for de-isolation of COVID-19 patients. Available at: https://www.ams.edu.sg/view-pdf.aspx?file=media%5c5557_fi_20.pdf&ofile=Position+Statement+from+the+NCID+%26+Chapter+of+ID++-+Period+of+Infectivity+Position+Statement+23-5-20.pdf

16. Nalla AK, Casto AM, Huang M-L, et al. Comparative performance of SARS-CoV-2 detection assays using seven different primer-probe sets and one assay kit. J Clin Microbiol 2020; 58:e00557–20. doi: 10.1128/JCM.00557-20.

17. Arons MM, Hatfield KM, Reddy SC, et al Presymptomatic SARS-CoV-2 Infections and Transmission in a Skilled Nursing Facility. New Engl J Med. 2020;382(22):2081–90.

18. Yang G, Erdman DE, Kodani M et al. Comparison of commercial systems for extraction of nucleic acids from DNA/RNA respiratory pathogens. J Virol Methods. 2011 Jan;171(1) 195–199. doi:10.1016/j.jviromet.2010.10.024. PMID: 21034773; PMCID: PMC7112907.

